# Plasma Proteomic Correlates of Cognitive Function in Older Adults From Central Africa

**DOI:** 10.64898/2026.10.22.26366214

**Authors:** Saranya Sundaram Patel, Peng Zhang, Jingjing Yang, Morgen Crosby, Caterina Obenauf, Guy Gikelekele, Emmanuel Epenge, Japhet Magolu Potshi, Topina Tomadia, Francine Manyonga Sabowa, Joseph Phuati Tsangu, Samuel Mampunza, Lelo Mananga, Xuemei Zeng, Thomas K. Karikari, Alden L. Gross, Jean Ikanga

**Affiliations:** Emory University School of Medicine, Department of Rehabilitation Medicine, Atlanta, GA 30322, USA; OneRehab, Dallas, Texas, USA; Emory University, School of Medicine, Department of Human Genetics and Biomedical Informatics, Atlanta, GA 30322, USA; University of Tennessee, Department of Psychology and Neuroscience, Knoxville, TN, USA; University of Kinshasa and Catholic University of Congo, School of Medicine, Kinshasa, Department of Psychiatry, B.P. 7463 Kinshasa I, Democratic Republic of Congo; University of Kinshasa, Department of Neurology, Kinshasa, B.P. 7463 Kinshasa I, Democratic Republic of Congo; Memory Clinic of Kinshasa, Kinshasa, B.P. 7463 Kinshasa I, Democratic Republic of Congo; University of Kinshasa, Department of Anesthesiology and Reanimation, B.P. 7463 Kinshasa, Democratic Republic of Congo; University of Kinshasa and Université Protestante au Congo, Department of Psychiatry, B.P. 7463 Kinshasa I, Democratic Republic of Congo; Department of Psychiatry, University of Pittsburgh, Pittsburgh, PA, USA; Alzheimer’s Disease Research Center, University of Pittsburgh, Pittsburgh, PA, USA; Biofluid Biomarker Laboratory, Western Psychiatric Hospital, University of Pittsburgh Medical Center, Pittsburgh, PA, USA; Clinical and Translational Science Institute, University of Pittsburgh, Pittsburgh, PA, USA; Department of Epidemiology, Johns Hopkins Bloomberg School of Public Health, Baltimore, Maryland, USA

**Keywords:** plasma proteomics, cognition, neurofilament light, neprilysin, MME, p-tau217, sub-Saharan Africa, Democratic Republic of Congo

## Abstract

**Key Points:** *Question:* Which candidate plasma proteins are associated with cognitive performance across multiple domains in older adults from the Democratic Republic of Congo?

*Findings:* In this cross-sectional study of 779 participants, higher MME was associated with better language, global cognition, and delayed memory; higher NEFL was associated with poorer global cognition and delayed memory; and ENO2 was associated with delayed memory. No p-tau217-based measure remained significant after within-outcome multiple-testing correction.

*Meaning:* Plasma markers most closely associated with cognitive phenotype may differ from biomarkers optimized for Alzheimer pathology detection, underscoring the need for population-specific biomarker evaluation.

**Importance:** Blood-based proteomic biomarkers may support scalable dementia research, but their associations with cognitive phenotype remain poorly characterized in sub-Saharan African populations.

**Objective:** To examine associations between selected plasma proteins and multidomain cognitive performance in older adults in the Democratic Republic of Congo.

**Design:** Cross-sectional secondary analysis of baseline data from the Étude du Vieillissement Cognitif et de Démence en République Démocratique du Congo (EVCD-RDC), with participant recruitment and data collection from July 2024 through September 2025.

**Setting:** Community-based prospective cohort in Kinshasa, Democratic Republic of Congo.

**Participants:** The analytic sample included 779 participants: 143 cognitively unimpaired, 218 with mild cognitive impairment, 203 with Alzheimer disease, and 215 with other dementia. Median age was 73 years (IQR, 70-78); 495 participants (63.5%) were female and 284 (36.5%) were male.

**Exposures:** Plasma abundance of 10 candidate proteins measured using the NULISAseq CNS Disease Panel 120. Candidates were selected from an earlier panel-wide analysis based on cognitively unimpaired vs Alzheimer disease discrimination and false-discovery-rate–adjusted significance.

**Main Outcomes and Measures:** Seven cognitive outcomes spanning global cognition, language, working memory, learning, delayed memory, mental abstraction, and problem solving. Separate linear regression models adjusted for age, sex, current smoking, education, and estimated glomerular filtration rate. Benjamini-Hochberg false-discovery-rate correction was applied within each cognitive outcome across the 10 proteins.

**Results:** Eighteen protein-cognition associations were nominally significant, and 6 remained significant after correction. Higher MME was associated with better language (β=0.143; 95% CI, 0.074-0.212), global cognition (β=0.225; 95% CI, 0.071-0.379), and delayed memory (β=0.312; 95% CI, 0.106-0.519). Higher NEFL was associated with poorer global cognition (β=-0.700; 95% CI, −1.080 to −0.319) and delayed memory (β=-0.773; 95% CI, −1.283 to −0.263). Higher ENO2 was associated with better delayed memory (β=0.560; 95% CI, 0.184-0.937). No p-tau217-based measure remained significant after correction.

**Conclusions and Relevance:** MME and NEFL showed the most consistent associations with cognitive performance, whereas p-tau217-based measures did not survive multiple-testing correction. Biomarkers optimized for Alzheimer pathology detection may differ from those most closely associated with cross-sectional cognitive phenotype, supporting population-specific evaluation of plasma proteomic markers.

## INTRODUCTION

Dementia prevalence is increasing rapidly worldwide, with much of the projected growth expected in low- and middle-income countries.^1–3^ Despite this burden, contemporary biomarker research remains disproportionately based on cohorts from high-income countries and populations of European ancestry. This imbalance is particularly important in sub-Saharan Africa, where vascular and metabolic risk, environmental exposures, educational opportunity, linguistic context, and access to specialist care may differ from the settings in which most biomarker models were developed. Emerging proteomic studies in African populations have identified both shared and population-specific Alzheimer disease (AD)-related signals, underscoring the need to evaluate biomarker–phenotype relationships directly in African cohorts rather than assume transportability.^4^

Blood-based biomarkers provide a scalable approach to characterizing neurodegenerative disease biology. Plasma phosphorylated tau, amyloid-β measures, neurofilament light (NfL), and multiplex proteomic profiles have demonstrated value for disease detection, biological classification, and prognosis.^5–7^ However, much of this work has focused on diagnostic or pathological classification rather than the relationship between circulating proteins and specific dimensions of cognitive performance. Recent studies using the NULISA platform have shown that plasma proteomic profiles capture biological information beyond conventional AD biomarkers. Gills and colleagues identified proteins associated with global cognition and executive function,^8^ while independent NULISA studies demonstrated strong p-tau217 performance for amyloid-related pathology together with inflammatory, vascular, synaptic, and other proteomic signals.^9–11^ Broader plasma proteomic studies have likewise identified multianalyte signatures associated with cognitive decline and incident dementia.^12^ These findings suggest that proteins most useful for detecting AD pathology may not necessarily be those most closely associated with cross-sectional cognitive phenotype. Comparable domain-level proteomic data from Central African populations remain sparse.

The Étude du Vieillissement Cognitif et de Démence en République Démocratique du Congo (EVCD-RDC) is a prospective cohort in Kinshasa designed to study cognitive aging and dementia using culturally adapted clinical, neuropsychological, and biomarker assessments.^13^ Prior work in this cohort demonstrated associations between plasma AD-related biomarkers and culturally adapted measures of cognition.^14^ The present study extends that work using high-sensitivity multiplex proteomics. Ten candidate plasma proteins were carried forward from an earlier panel-wide NULISAseq analysis based on their ability to discriminate cognitively unimpaired participants from those with AD and on false-discovery-rate–adjusted statistical significance; selection was independent of the cognition analyses reported here. These candidates included proteins reflecting amyloid and tau biology, neuroaxonal injury, neuronal metabolism, inflammation, and extracellular-matrix processes, including membrane metalloendopeptidase (MME/neprilysin), NfL, and p-tau217–based measures.^15–20^

We examined associations between these 10 candidate plasma proteins and 7 cognitive outcomes spanning global cognition, language, working memory, learning, delayed memory, mental abstraction, and problem solving. Our objective was to identify protein-cognition associations that remained significant after correction within cognitive outcomes and to determine whether proteins selected for diagnostic discrimination showed similar relationships with cognitive phenotype. By focusing on multidomain cognition rather than diagnostic classification, this study aimed to clarify how circulating proteomic signals relate to cognitive function in an older Central African population that remains substantially underrepresented in dementia biomarker research.

## METHODS

### Study design and participants

This cross-sectional secondary analysis used baseline data from the Étude du Vieillissement Cognitif et de Démence en République Démocratique du Congo (EVCD-RDC), a prospective longitudinal cohort based in Kinshasa, Democratic Republic of Congo. Participant recruitment and baseline data collection occurred from July 2024 through September 2025.^13^ Parent-cohort eligibility required Congolese citizenship and residence in the DRC, age 65 years or older, fluency in French or Lingala, ability to provide informed consent independently or through an authorized representative, reliable contact information including a caregiver or informant, and willingness to participate longitudinally. Individuals with major sensory impairment or neurodevelopmental, psychiatric, or neurological conditions expected to compromise central nervous system assessment were excluded. No additional age restriction was applied for the present analysis. Of 1,872 individuals screened for the parent cohort, 1,260 completed baseline evaluation following informed consent, 1,004 completed biospecimen collection, and 898 had completed multidisciplinary diagnostic adjudication. Of these, 119 did not meet one or more of the prespecified analytic requirements for the present study, including an eligible matched baseline clinical record with valid MoCA form status and a matched proteomic measurement meeting Passed or Warning quality-control criteria. The final analytic cohort therefore included 779 participants. The analytic cohort retained 779 participants with a recorded neurological diagnosis, an eligible matched baseline clinical record with valid MoCA form status, and a matched proteomic measurement assigned a Passed or Warning quality-control status. Participants classified as other dementia also had a recorded dementia subtype. The final cohort comprised 143 cognitively unimpaired (CU) participants, 218 with mild cognitive impairment (MCI), 203 with Alzheimer disease (AD), and 215 with other dementia (OD). Race and ethnicity were not collected as study variables. All participants met parent-cohort eligibility criteria requiring Congolese citizenship and residence in the Democratic Republic of Congo. The EVCD-RDC protocol was approved by the institutional review boards/ethics committees of the University of Kinshasa and Emory University. All participants provided written informed consent before participation. Study procedures were conducted in accordance with applicable ethical standards and the Declaration of Helsinki.

### Cognitive assessment

Participants completed cognitive assessments in French, Lingala, or both according to language proficiency and preference. Measures were culturally adapted, translated using forward-backward procedures, and piloted in the Congolese context.^13^˒^21^˒^22^ Seven outcomes were examined: global cognition (MoCA total score), working memory (African Market Test working-memory score), language (Image Naming and Recognition Test unprompted naming/recognition score), learning (combined third-trial scores from the African List Memory Test and African Visuospatial Memory Test), delayed memory (corresponding verbal and visuospatial delayed-recall scores), mental abstraction (African Proverbs Test total score), and problem solving (African Card Game Test total won). Mental abstraction and problem solving were analyzed separately because the measures have different raw-score ranges.

### Biospecimen collection and plasma proteomics

Blood was collected at Centre Médical de Kinshasa, processed from EDTA plasma, aliquoted, and stored at −80 °C. Plasma proteins were measured using the NULISAseq CNS Disease Panel 120 (Alamar Biosciences) on the ARGO HT platform according to published protocols and manufacturer specifications.⁹^-^¹¹ The panel quantified 131 protein biomarkers using DNA-barcoded antibody pairs and next-generation sequencing. Sequencing counts were normalized to an internal mCherry standard, harmonized using interplate controls, and processed by the assay pipeline to generate NULISA Protein Quantification (NPQ) values.

Samples were analyzed across 10 plates. Median intra- and interplate coefficients of variation for vendor controls were 6.3% and 5.8%, respectively; the median interpolated interplate coefficient of variation for in-house pooled quality-control samples was approximately 11%. For the present analyses, stored assay-derived NPQ values were analyzed on the scale provided in the dataset without additional transformation. Measurements with Passed or Warning quality-control status were retained.

### Candidate protein measures

Ten candidate protein measures were examined: MME, NEFL, SMOC1, brain-derived p-tau217/Aβ42, MDH1, IL6R, IFNG, brain-derived p-tau217, ENO2, and p-tau217. The brain-derived p-tau217/Aβ42 ratio was calculated from the corresponding stored NPQ values. Candidates were selected before the present cognition analyses from an earlier panel-wide NULISAseq analysis based jointly on CU-vs-AD discrimination and FDR-adjusted statistical significance. Selection was therefore independent of the protein-cognition associations examined here.

### Covariates

Models adjusted for age, sex, current smoking, years of education, and estimated glomerular filtration rate (eGFR). These covariates were selected to account for demographic, educational, behavioral, and renal factors that could influence cognitive performance or circulating protein abundance. Current smoking was coded yes/no. eGFR was calculated using the MDRD equation used in the EVCD-RDC cohort: eGFR = 175 × (serum creatinine)^−1.154 × (age)^−0.203 × (0.742 if female).¹³ Sex was recorded as part of the baseline demographic assessment and analyzed as recorded in the EVCD-RDC dataset.

### Statistical analysis

No a priori sample size calculation was performed; all participants who met the prespecified analytic eligibility criteria were included. Each protein-cognition pair was evaluated in a separate multivariable linear regression model adjusted for age, sex, current smoking, years of education, and eGFR. Seven cognitive outcomes and 10 proteins yielded 70 models. Analyses used complete cases for the outcome, protein measure, and covariates; no imputation was performed. Model-specific sample sizes were 667 for global cognition, 330 for working memory, 678 for language, 680 for learning, 680 for delayed memory, 553 for mental abstraction, and 680 for problem solving; variation reflected outcome- and covariate-specific missingness. All tests were 2-sided. Cognitive outcomes and protein NPQ variables were analyzed on their original stored scales; standardized coefficients were additionally calculated for descriptive comparison. We report unstandardized regression coefficients, 95% CIs, and nominal P values.

To address multiple testing, P values were adjusted using the Benjamini-Hochberg false-discovery-rate procedure separately within each cognitive outcome across its 10 candidate proteins. Associations with q<.05 were considered statistically significant; associations with nominal P<.05 that did not survive correction were considered exploratory. Analyses were conducted using R version 4.5.3 (R Foundation for Statistical Computing). Data management and visualization used tidyverse version 2.0.0 and patchwork version 1.3.2; regression diagnostics used lmtest version 0.9.40 and car version 3.1.5.

For descriptive comparisons across CU, MCI, AD, and other dementia groups, continuous variables were compared using Kruskal-Wallis tests and categorical variables using Pearson χ² tests. BH-FDR correction was applied separately within the demographic, clinical/cognitive, and protein sections of Table 1.

**Table 1.**
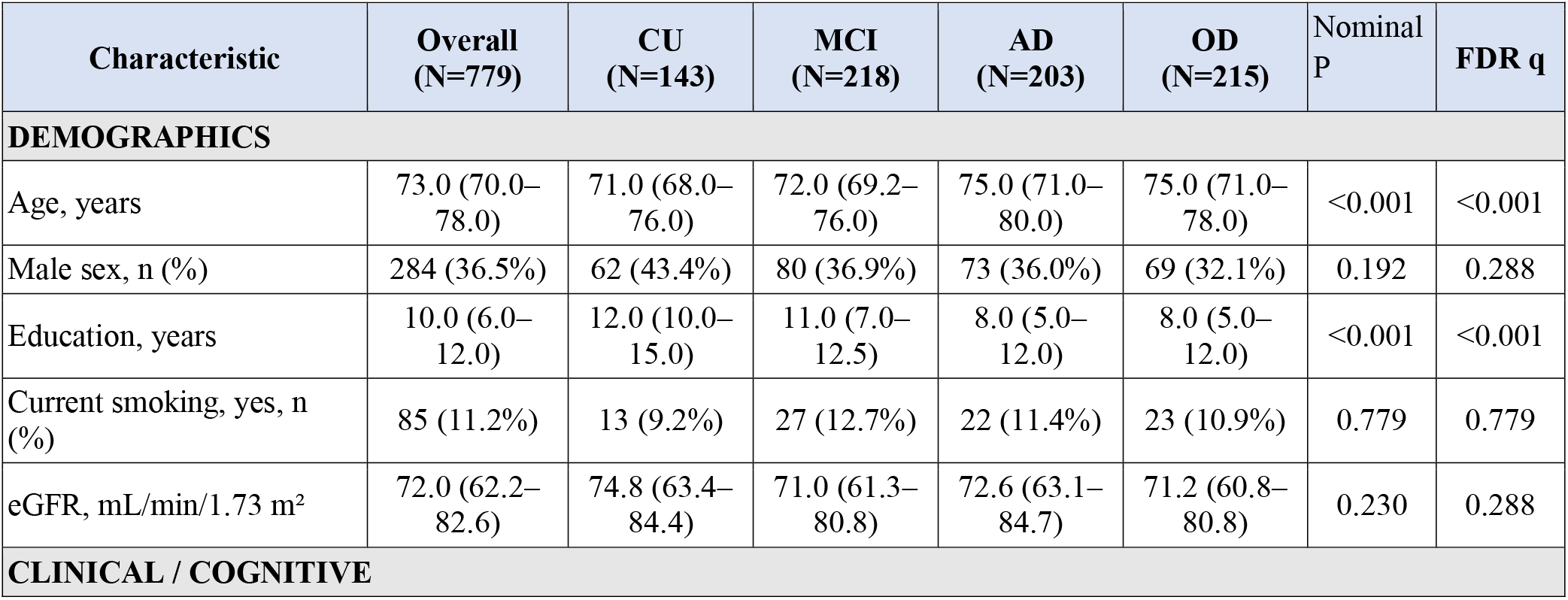
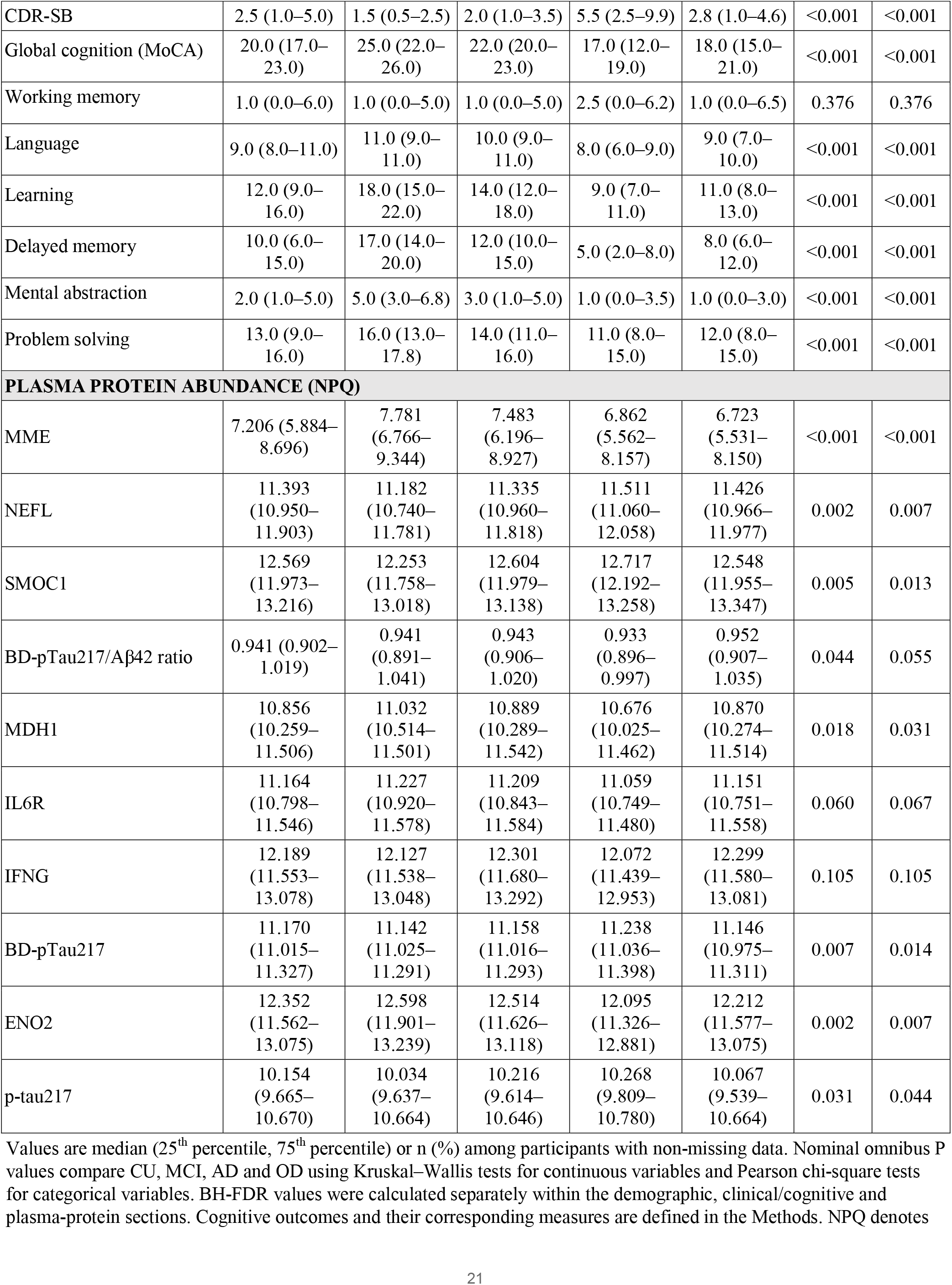
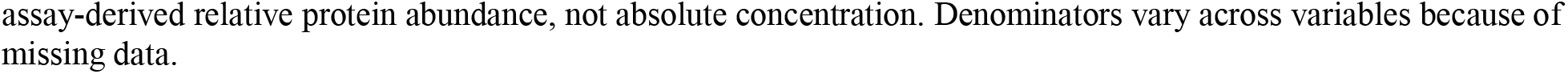
Participant characteristics, cognitive performance, and candidate plasma protein abundance stratified by neurological status.

| Characteristic | Overall<br>(N=779) | CU<br>(N=143) | MCI<br>(N=218) | AD<br>(N=203) | OD<br>(N=215) | Nominal<br>P | FDR q |
| --- | --- | --- | --- | --- | --- | --- | --- |
| **DEMOGRAPHICS** |  |  |  |  |  |  |  |
| Age, years | 73.0 (70.0–78.0) | 71.0 (68.0–76.0) | 72.0 (69.2–76.0) | 75.0 (71.0–80.0) | 75.0 (71.0–78.0) | <0.001 | <0.001 |
| Male sex, n (%) | 284 (36.5%) | 62 (43.4%) | 80 (36.9%) | 73 (36.0%) | 69 (32.1%) | 0.192 | 0.288 |
| Education, years | 10.0 (6.0–12.0) | 12.0 (10.0–15.0) | 11.0 (7.0–12.5) | 8.0 (5.0–12.0) | 8.0 (5.0–12.0) | <0.001 | <0.001 |
| Current smoking, yes, n (%) | 85 (11.2%) | 13 (9.2%) | 27 (12.7%) | 22 (11.4%) | 23 (10.9%) | 0.779 | 0.779 |
| eGFR, mL/min/1.73 m<sup>2</sup> | 72.0 (62.2–82.6) | 74.8 (63.4–84.4) | 71.0 (61.3–80.8) | 72.6 (63.1–84.7) | 71.2 (60.8–80.8) | 0.230 | 0.288 |
| **CLINICAL / COGNITIVE** |  |  |  |  |  |  |  |
| CDR-SB | 2.5 (1.0–5.0) | 1.5 (0.5–2.5) | 2.0 (1.0–3.5) | 5.5 (2.5–9.9) | 2.8 (1.0–4.6) | <0.001 | <0.001 |
| Global cognition (MoCA) | 20.0 (17.0–23.0) | 25.0 (22.0–26.0) | 22.0 (20.0–23.0) | 17.0 (12.0–19.0) | 18.0 (15.0–21.0) | <0.001 | <0.001 |
| Working memory | 1.0 (0.0–6.0) | 1.0 (0.0–5.0) | 1.0 (0.0–5.0) | 2.5 (0.0–6.2) | 1.0 (0.0–6.5) | 0.376 | 0.376 |
| Language | 9.0 (8.0–11.0) | 11.0 (9.0–11.0) | 10.0 (9.0–11.0) | 8.0 (6.0–9.0) | 9.0 (7.0–10.0) | <0.001 | <0.001 |
| Learning | 12.0 (9.0–16.0) | 18.0 (15.0–22.0) | 14.0 (12.0–18.0) | 9.0 (7.0–11.0) | 11.0 (8.0–13.0) | <0.001 | <0.001 |
| Delayed memory | 10.0 (6.0–15.0) | 17.0 (14.0–20.0) | 12.0 (10.0–15.0) | 5.0 (2.0–8.0) | 8.0 (6.0–12.0) | <0.001 | <0.001 |
| Mental abstraction | 2.0 (1.0–5.0) | 5.0 (3.0–6.8) | 3.0 (1.0–5.0) | 1.0 (0.0–3.5) | 1.0 (0.0–3.0) | <0.001 | <0.001 |
| Problem solving | 13.0 (9.0–16.0) | 16.0 (13.0–17.8) | 14.0 (11.0–16.0) | 11.0 (8.0–15.0) | 12.0 (8.0–15.0) | <0.001 | <0.001 |
| **PLASMA PROTEIN ABUNDANCE (NPQ)** |  |  |  |  |  |  |  |
| MME | 7.206 (5.884–8.696) | 7.781 (6.766–9.344) | 7.483 (6.196–8.927) | 6.862 (5.562–8.157) | 6.723 (5.531–8.150) | <0.001 | <0.001 |
| NEFL | 11.393 (10.950–11.903) | 11.182 (10.740–11.781) | 11.335 (10.960–11.818) | 11.511 (11.060–12.058) | 11.426 (10.966–11.977) | 0.002 | 0.007 |
| SMOC1 | 12.569 (11.973–13.216) | 12.253 (11.758–13.018) | 12.604 (11.979–13.138) | 12.717 (12.192–13.258) | 12.548 (11.955–13.347) | 0.005 | 0.013 |
| BD-pTau217/Aβ42 ratio | 0.941 (0.902–1.019) | 0.941 (0.891–1.041) | 0.943 (0.906–1.020) | 0.933 (0.896–0.997) | 0.952 (0.907–1.035) | 0.044 | 0.055 |
| MDH1 | 10.856 (10.259–11.506) | 11.032 (10.514–11.501) | 10.889 (10.289–11.542) | 10.676 (10.025–11.462) | 10.870 (10.274–11.514) | 0.018 | 0.031 |
| IL6R | 11.164 (10.798–11.546) | 11.227 (10.920–11.578) | 11.209 (10.843–11.584) | 11.059 (10.749–11.480) | 11.151 (10.751–11.558) | 0.060 | 0.067 |
| IFNG | 12.189 (11.553–13.078) | 12.127 (11.538–13.048) | 12.301 (11.680–13.292) | 12.072 (11.439–12.953) | 12.299 (11.580–13.081) | 0.105 | 0.105 |
| BD-pTau217 | 11.170 (11.015–11.327) | 11.142 (11.025–11.291) | 11.158 (11.016–11.293) | 11.238 (11.036–11.398) | 11.146 (10.975–11.311) | 0.007 | 0.014 |
| ENO2 | 12.352 (11.562–13.075) | 12.598 (11.901–13.239) | 12.514 (11.626–13.118) | 12.095 (11.326–12.881) | 12.212 (11.577–13.075) | 0.002 | 0.007 |
| p-tau217 | 10.154 (9.665–10.670) | 10.034 (9.637–10.664) | 10.216 (9.614–10.646) | 10.268 (9.809–10.780) | 10.067 (9.539–10.664) | 0.031 | 0.044 |

Values are median (25<sup>th</sup> percentile, 75<sup>th</sup> percentile) or n (%) among participants with non-missing data. Nominal omnibus P values compare CU, MCI, AD and OD using Kruskal–Wallis tests for continuous variables and Pearson chi-square tests for categorical variables. BH-FDR values were calculated separately within the demographic, clinical/cognitive and plasma-protein sections. Cognitive outcomes and their corresponding measures are defined in the Methods. NPQ denotes
assay-derived relative protein abundance, not absolute concentration. Denominators vary across variables because of missing data.

Model assumptions were evaluated using residual Q-Q plots, residual-versus-fitted plots, Breusch-Pagan tests, and variance-inflation factors.^23^ Diagnostics informed interpretation but did not result in transformation or exclusion of variables.

## RESULTS

### Participant characteristics and analytic sample

The analytic cohort included 779 participants: 143 cognitively unimpaired (CU), 218 with mild cognitive impairment (MCI), 203 with Alzheimer disease (AD), and 215 with other dementia (OD). Median age was 73 years (IQR, 70-78), 284 participants (36.5%) were male, and median educational attainment was 10 years (IQR, 6-12). Neurological-status groups differed in age and education after section-wise FDR correction (both q<.001), but not in sex, current smoking, or eGFR. Global cognition, CDR-SB, language, learning, delayed memory, mental abstraction, and problem solving differed across groups (all q<.001), whereas working memory did not (q=.376; Table 1).

### Protein abundance differed across neurological-status groups

Seven of the 10 candidate proteins differed across neurological-status groups after section-wise FDR correction: MME, NEFL, SMOC1, MDH1, brain-derived p-tau217, ENO2, and p-tau217 (Table 1). Descriptively, MME and ENO2 were higher in CU than in AD, whereas NEFL was higher in AD. Because these were omnibus comparisons, these patterns should not be interpreted as pairwise group differences.

### Protein-cognition associations after multiple-testing correction

Across the 70 adjusted models, 18 protein–cognition associations had nominal P<.05, and 6 remained significant after BH-FDR correction within cognitive outcomes (Figure 1A). Higher MME was associated with better language performance (β=0.143; 95% CI, 0.074-0.212; q<.001), global cognition (β=0.225; 95% CI, 0.071-0.379; q=.021), and delayed memory (β=0.312; 95% CI, 0.106-0.519; q=.012). Higher NEFL was associated with poorer global cognition (β=-0.700; 95% CI, −1.080 to −0.319; q=.003) and delayed memory (β=-0.773; 95% CI, −1.283 to −0.263; q=.012). Higher ENO2 was associated with better delayed memory (β=0.560; 95% CI, 0.184-0.937; q=.012).

**Figure 1.**
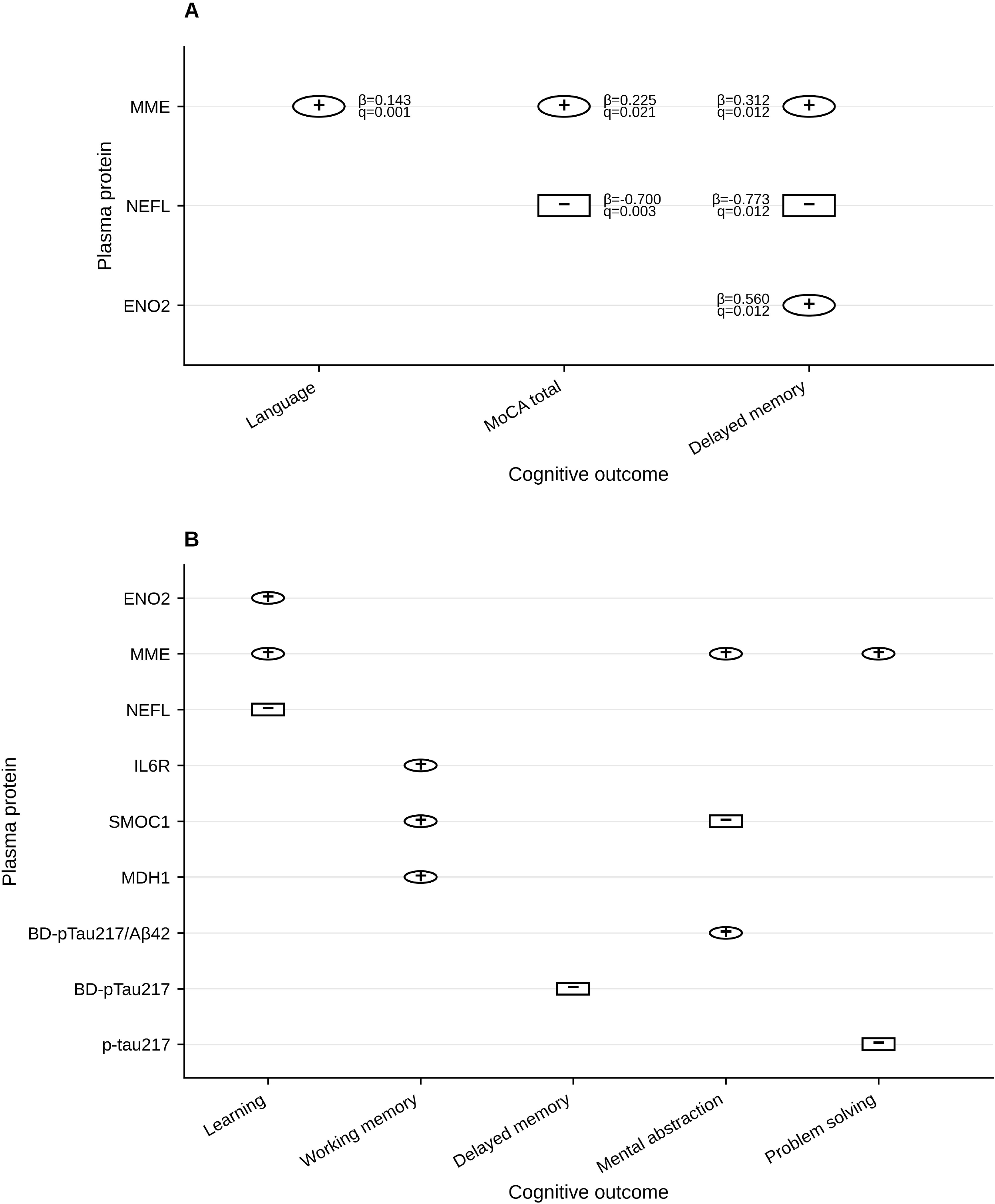
Protein-Cognition Associations by Multiple-Testing Significance. A, Associations that remained significant after within-outcome Benjamini-Hochberg false discovery rate (FDR) correction. Labels indicate unstandardized regression coefficients (β) and FDR-adjusted q values. B, Associations with nominal P<.05 that did not remain significant after within-outcome FDR correction. Circles indicate positive associations, and squares indicate inverse associations. Each association was estimated in a separate multivariable linear regression model adjusted for age, sex, current smoking, years of education, and estimated glomerular filtration rate. FDR correction was applied separately within each cognitive outcome across the 10 candidate proteins. Abbreviations: BD-pTau217, brain-derived phosphorylated tau 217; ENO2, enolase 2; IL6R, interleukin 6 receptor; MDH1, malate dehydrogenase 1; MME, membrane metalloendopeptidase/neprilysin; NEFL, neurofilament light; SMOC1, SPARC-related modular calcium-binding protein 1.

### Nominal associations not retained after within-outcome multiple-testing correction

Twelve additional associations had nominal P<.05 but did not survive within-outcome FDR correction (Figure 1B). Each of the 3 p-tau217-based measures showed at least 1 nominal domain-specific association, but none remained significant after correction. The full pattern of adjusted protein–cognition associations is shown in Figure 2.

**Figure 2.**
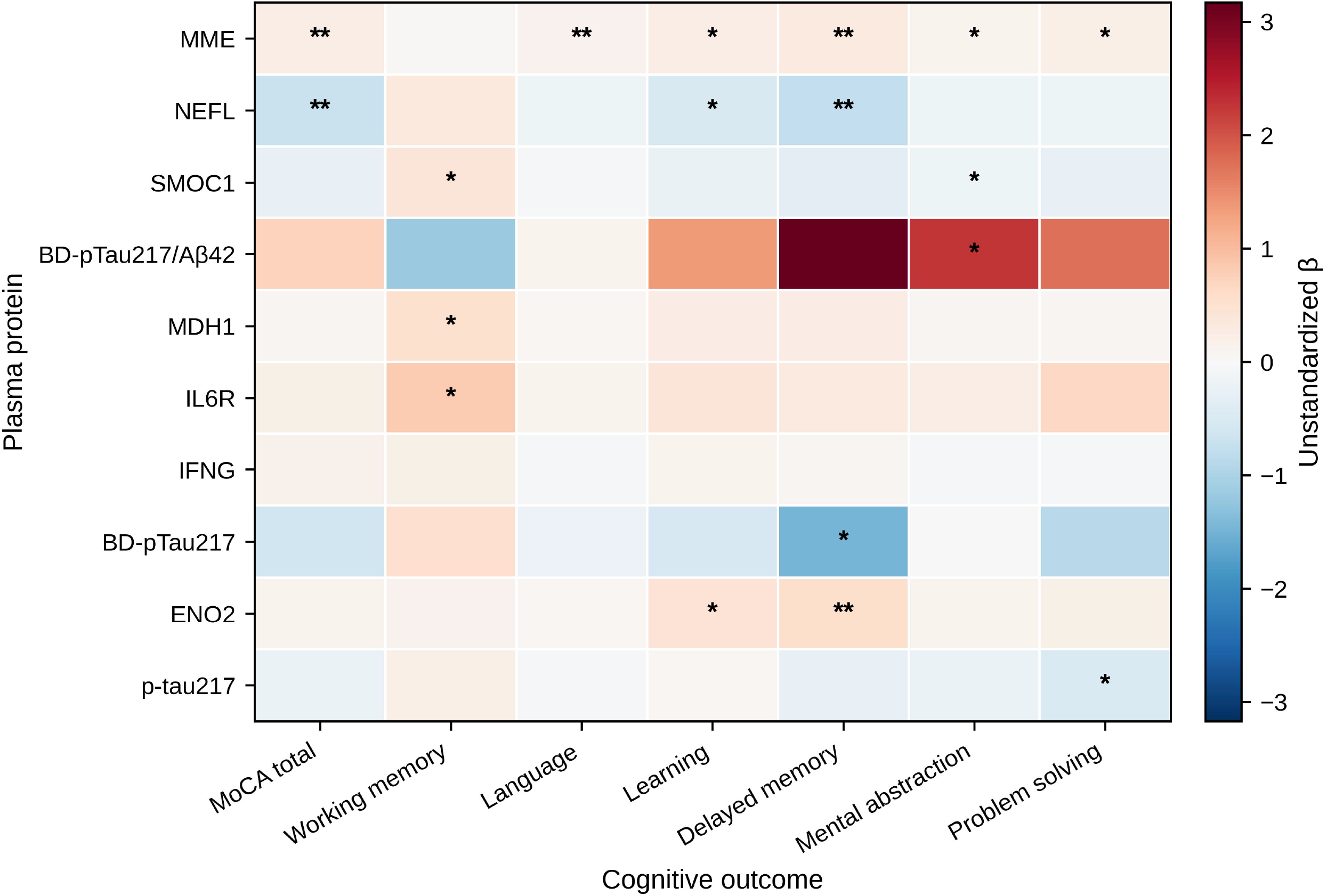
Adjusted Protein-Cognition Association Map. Heat map showing unstandardized regression coefficients (β) from 70 separate multivariable linear regression models relating 10 candidate plasma proteins to 7 cognitive outcomes. Models were adjusted for age, sex, current smoking, years of education, and estimated glomerular filtration rate. Red indicates positive associations and blue indicates inverse associations; greater color intensity indicates a larger absolute unstandardized coefficient. Because proteins and cognitive outcomes were analyzed on different scales, color intensity should not be interpreted as directly comparable effect magnitude across all cells. A single asterisk indicates nominal P<.05; a double asterisk indicates q<.05 after Benjamini-Hochberg FDR correction applied separately within each cognitive outcome across the 10 candidate proteins. Abbreviations: BD-pTau217, brain-derived phosphorylated tau 217; ENO2, enolase 2; IFNG, interferon gamma; IL6R, interleukin 6 receptor; MDH1, malate dehydrogenase 1; MME, membrane metalloendopeptidase/neprilysin; NEFL, neurofilament light; SMOC1, SPARC-related modular calcium-binding protein 1.

## DISCUSSION

In this cross-sectional study of 779 older adults from the Democratic Republic of Congo, plasma proteomic measures showed distinct associations with cognitive performance across multiple domains. Six associations remained significant after BH-FDR correction within cognitive outcomes. Higher MME was associated with better language, global cognition, and delayed memory, whereas higher NEFL was associated with poorer global cognition and delayed memory; higher ENO2 was additionally associated with better delayed memory. In contrast, none of the p-tau217–based measures remained significantly associated with cognition after multiple-testing correction. These findings highlight an important distinction between biomarkers that discriminate clinical or pathological states and those most closely associated with cross-sectional cognitive phenotype.

The opposing associations of MME and NEFL with cognition are notable because these proteins reflect different biological processes. NEFL is an established marker of neuroaxonal injury, and its inverse associations with global cognition and delayed memory are consistent with prior studies linking higher circulating NfL to greater neurodegenerative burden and poorer cognitive outcomes.^19^˒^20^ The descriptive neurological-status comparisons were directionally concordant, with NEFL lowest in cognitively unimpaired participants and highest in those with AD, although these omnibus comparisons should not be interpreted as evidence of pairwise or graded disease-stage differences.

MME demonstrated the broadest pattern of cognitive associations, spanning language, global cognition, and delayed memory, with additional nominal associations in other domains. Neprilysin is involved in amyloid-β degradation,^15–17^ and pharmacologic neprilysin inhibition can alter circulating amyloid measures.^18^ However, MME is expressed in multiple peripheral tissues and participates in vascular, metabolic, and inflammatory signaling. Plasma MME abundance therefore should not be interpreted as a direct measure of cerebral amyloid clearance or as evidence that greater neprilysin abundance is neuroprotective. Rather, the present results identify MME as a circulating protein associated with cognitive phenotype and support further investigation of its biological and clinical relevance.

These findings are consistent with a broader literature suggesting that cognition reflects multiple circulating biological pathways rather than a single disease-specific biomarker. Prior NULISA studies have identified proteins associated with cognition and neurodegenerative phenotypes while also demonstrating strong p-tau217 performance for amyloid-related pathology.^8–11^ Broader proteomic studies have similarly identified multianalyte signatures associated with cognitive decline and incident dementia, including externally replicated associations involving NEFL.^12^ MME has not consistently emerged as a cognition-associated protein in prior plasma proteomic studies, making replication of the present multidomain associations particularly important. Importantly, these results do not establish MME as a diagnostic biomarker; instead, they suggest that a protein may be related to cognitive phenotype even when its role in diagnostic classification remains uncertain.

The positive association between ENO2 and delayed memory warrants particular caution. ENO2 encodes neuron-specific enolase, a glycolytic enzyme that may be released during neuronal injury, making the observed direction difficult to reconcile with a simple injury-marker model.^24^ Nevertheless, ENO2 abundance was descriptively higher in cognitively unimpaired participants and lower in participants with AD, a pattern directionally consistent with the continuous cognitive association. Nonlinear relationships, disease-stage effects, peripheral contributions, residual confounding, or assay-specific characteristics could contribute to this finding. Because the present analyses were cross-sectional, replication and longitudinal evaluation are needed before assigning mechanistic significance to the ENO2 association.

The p-tau217 findings also require careful interpretation. Although brain-derived p-tau217 and p-tau217 differed across neurological-status groups, none of the 3 p-tau217-based measures showed cognitive associations that survived within-outcome FDR correction. This does not suggest that p-tau217 lacks value as an Alzheimer disease biomarker. Rather, accumulating evidence indicates that p-tau217 is highly informative for AD pathology while its distributions and associations with cognition may vary across populations and cognitive domains.^25–27^ Prior NULISA studies have demonstrated strong p-tau217 performance for amyloid-related classification,^9–11^ whereas diverse-cohort studies have reported variation in p-tau217 concentrations and cognition associations across racial and ethnic groups.^25^˒^26^ Conversely, p-tau217/Aβ42 has demonstrated high amyloid-PET classification accuracy across Black and White participants.^27^ Taken together, these findings emphasize that pathological classification and cross-sectional association with cognition represent related but distinct biomarker properties.

This study combines high-sensitivity plasma proteomics with multidomain cognitive phenotyping in a Central African cohort that remains substantially underrepresented in contemporary dementia biomarker research. Cognitive outcomes incorporated measures developed or adapted for use in African populations, reducing reliance on instruments designed exclusively for high-income Western settings.^13^˒^14^˒^21^˒^28^ The analysis also accounted for education, smoking, and renal function and controlled multiple testing within each cognitive outcome.

Several limitations should be considered. First, the cross-sectional design precludes conclusions regarding temporal relationships, cognitive decline, or causality. Second, the cohort was clinically heterogeneous, and models did not adjust for diagnosis or several potentially relevant cardiometabolic and genetic factors, including diabetes, body mass index, hypertension, medication use, and APOE status. Third, outcome-specific missingness resulted in different analytic samples across cognitive domains, particularly for working memory. Fourth, the 10 proteins were selected from an earlier CU-vs-AD analysis based on diagnostic discrimination and FDR-adjusted significance rather than through a proteome-wide screen for cognition; proteins associated with cognition through other pathways may therefore have been missed. Fifth, models used the original outcome and NPQ scales, and residual diagnostics indicated nonconstant variance for some outcome families. Circulating protein abundance also may not reflect brain expression, enzymatic activity, or tissue of origin. Finally, recruitment from Kinshasa may limit generalizability to other Congolese or African populations.

Longitudinal follow-up within EVCD-RDC should determine whether MME, NEFL, ENO2, and other candidate proteins predict domain-specific cognitive decline and whether the observed associations persist after accounting for clinical diagnosis, cardiometabolic factors, established AD biomarkers, and other biological measures. Replication across African cohorts will be important for distinguishing broadly transportable biomarker relationships from population- or context-specific associations.

In conclusion, plasma MME and NEFL showed the most consistent associations with multidomain cognitive performance in older Congolese adults, whereas no p-tau217–based measure remained significant after within-outcome multiple-testing correction. These findings underscore that biomarkers optimized for Alzheimer pathology detection may differ from those most closely associated with cross-sectional cognitive phenotype. Population-specific evaluation of plasma proteomic markers will be important as blood-based biomarker frameworks expand globally.

## Author Contributions

Drs. Ikanga and Yang had full access to all of the data in the study and take responsibility for the integrity of the data and the accuracy of the data analysis.

**Concept and design**: Patel, Zhang, Yang, Karikari, Ikanga.

**Acquisition, analysis, or interpretation of data**: All authors.

**Drafting of the manuscript**: Patel and Ikanga.

**Critical revision of the manuscript for important intellectual content**: All authors.

**Statistical analysis**: Zhang, Yang.

**Obtained funding**: Ikanga and Gross.

**Administrative, technical, or material support**: Gikelekele, Epenge, Potshi, Tomadia, Sabowa, Tsangu, Mampunza, Mananga, Zeng, Karikari.

**Supervision**: Yang, Karikari, Ikanga.

## Conflict of Interest Disclosures

None reported.

## Funding/Support

This work was supported by an Alzheimer’s Association Research Grant, the Emory Goizueta Alzheimer’s Disease Research Center, and the National Institute on Aging of the National Institutes of Health (P30AG066511). This work was also supported by the National Center for Advancing Translational Sciences of the National Institutes of Health (UL1TR002378).

## Role of the Funder/Sponsor

The funders had no role in the design and conduct of the present study; collection, management, analysis, and interpretation of the data; preparation, review, or approval of the manuscript; or decision to submit the manuscript for publication.

## Additional Contributions

The authors thank the Centre Médical de Kinshasa for support with biospecimen collection, processing, and storage.

## Data Availability

All data produced in the present study are available upon reasonable request to the authors.

## Notes

### Competing Interest Statement

The authors have declared no competing interest.

### Author Declarations

The EVCD RDC protocol was approved by the institutional review boards/ethics committees of the University of Kinshasa and Emory University. All participants provided written informed consent before participation. Study procedures were conducted in accordance with applicable ethical standards and the Declaration of Helsinki.

